# Pilot implementation of ‘Outsourced Oxygen to the Bedside’ models in five countries: a mixed methods impact assessment

**DOI:** 10.64898/2026.02.21.26346705

**Authors:** Freddy Eric Kitutu, Celia Blaas, Pius Mukisa, Mattias Schedwin, Tim Baker, Ayobami A Bakare, Deepa Bishit, Elibariki Mkumbo, Jacquie Oliwa, Sophie Namasopo, Jacinta Nzinga, Maire Ruane, Aishat Adeniji, Michael Hawkes, Amarpreet Rai, Michael Njuguna, Hamish Graham, Carina King

## Abstract

**Background:** Medical oxygen is an essential medicine that is often unavailable for patients when they need it. We explored if ‘Outsourced Oxygen to the Bedside’ (O2B) pilots, where private providers deliver a package of services, were successful in ensuring reliable oxygen access at the patient bedside.

**Methods:** We conducted a sequential explanatory mixed-methods assessment of O2B pilots in Kenya, Nigeria, India, Tanzania, and Uganda from September 2024 – January 2025. A quantitative cross-sectional facility audit described facility contexts, tested equipment functionality and assessed healthcare worker (HCW) oxygen knowledge. Qualitative interviews with HCWs and managers explored experiences of O2B pilots.

**Results:** We studied 28 of the 80 facilities participating in the pilots, 179 HCWs completed the knowledge survey, and 59 qualitative interviews were conducted. In the audit, we found O2B provided oxygen equipment more functional and usable than non-O2B equipment: 49.0% vs 30.1% (p-value<0.001) for cylinders, 82.9% vs 20.3% (p-value<0.001) for concentrators, and 84.0% vs 70.0% (p-value=0.172) for pulse oximeters. Overall, 21.8% (39/179) of HCWs had received training from O2B providers, and their oxygen knowledge was slightly higher than those who had not (mean score 15.3/24 vs 13.9/24, p-value=0.002). Qualitative interviews highlighted positive changes in oxygen access and the ability to treat patients, but also mixed understandings of the O2B services being provided, and requests for additional services.

**Conclusion:** O2B pilots appear to improve medical oxygen access, with effective maintenance and repair services being a key mechanism. However, tailoring to local needs and remaining gaps in HCW capacity need to be addressed.

## Introduction

Every year, an estimated 374 million people require medical oxygen for acute medical conditions, and yet 60% of the world’s population do not have access to reliable, affordable, quality medical oxygen services.^1^ This access gap is most pronounced in sub-Saharan Africa and South Asia, where 91% and 78% of patients respectively, in need of oxygen for acute medical conditions do not receive it. Lack of functional and appropriate oxygen equipment, including pulse oximeters, is one reason driving this access gap, with smaller, rural, public facilities being most affected.^1^

Given medical oxygen is an essential medicine, used across multiple conditions and all age groups, improving oxygen systems is a cost-effective investment that would advance global progress against 8 of the 9 health-related Sustainable Development Goals targets.^2^ The COVID-19 pandemic resulted in large investments in oxygen equipment, but accompanied with limited funding for strengthening distribution systems, supply chains for spare parts, biomedical engineering and healthcare worker capacity.^1^ These elements of the oxygen system are recognized as critical weaknesses, meaning even when oxygen equipment is present, it is often non-functional and or interupted.^3-7^

‘Oxygen as a service’ (O2aaS) is an approach that aims to simplify the often-complicated oxygen supply chain, and therefore improve oxygen access for patients, by having a single provider responsible for providing a bundle of services – rather than just providing medical oxygen as a commodity. The combinations of equipment and services delivered by O2aaS can vary, depending on the target facilities, their oxygen needs and context, from large district level hub-and-spoke models,^8^ to smaller concentrator or cylinder-based models. ‘Outsourced oxygen to the bedside’ (O2B) models are conceptualized as a sub-set of O2aaS, in which a private entity charges a bundled price for medical oxygen delivery to the patient bedside, supplemented with various levels of equipment management, maintenance, and repair services.^9^ Further services, such as training, pulse oximeters and consumables may also be included.

While this approach is promising, some concerns have been raised around the adoption, scale up and sustainability of these models in low-resource contexts without donor support, and the potential to increase inequities in oxygen access.^9^ We therefore assessed five different O2B implementation pilots to determine if they were effective in delivering reliable medical oxygen access to the bedside and examined the mechanisms of effect from the perspective of service providers, in different health system contexts.

## Methods

### Study design

We conducted a sequential explanatory mixed-method evaluation of five O2B pilot implementation projects in Kenya, Nigeria, India, Tanzania, and Uganda. The pilots were initiated as part of the ‘Oxygen CoLab’ programme - a collaboration between Brink and DT Global, funded by the UK Foreign, Commonwealth and Development Office. The Oxygen CoLab selected the five O2B providers through an open competitive consultation and provided them with funding and technical support to deliver oxygen using a service-based model (**Supplementary Appendix 1**).

The research team conducted an independent assessment of the O2B pilots, using a quantitative cross-sectional health facility audit between September 2024 and January 2025, to describe facility contexts, presence and functionality of oxygen equipment, and healthcare worker (HCW) oxygen and pulse oximetry knowledge and skills. This was followed by semi-structured qualitative interviews between November 2024 and January 2025, to explore HCW, facility manager, and policy maker experiences of the O2B pilots. In this paper, we focus on the impact of the O2B pilots on access to reliable oxygen at the patient bedside, including availability of appropriate medical equipment and accessories, their functionality and HCW capacity. A separate publication addresses the potential for O2B models to be routinely adopted in the health systems.

### Settings

The study was conducted in Kenya, Nigeria, India, Tanzania, and Uganda. In Kenya, the Centre for Public Health and Development (CPHD) were the O2B provider, working in 12 facilities in urban Nairobi for their first phase of implementation. In Nigeria, HealthPort worked with 7 facilities in peri-urban Lagos and Ogun States, Southwest Nigeria. In India, Sanrai worked in 19 facilities in rural Uttar Pradesh. In Tanzania the O2B provider was FREO2 in 37 rural facilities across the four regions of Manyara, Arusha, Kilimanjaro and Pwani including Mafia Island. In Uganda, the Association for Health Innovation in Africa (AFHIA) worked with 5 facilities in rural Rukunguri and Kanungu districts, in the Western Region. The O2B pilots were implemented in public, private and faith-based facilities of different sizes and capacity across the sites (**Table 1**). A description of the different health system contexts and the O2B packages provided is presented in **Supplementary Appendix 1**.

**Table 1:** Description of the O2B pilot contexts.

|  | **Kenya** | **Nigeria** | **India** | **Tanzania** | **Uganda** |
| --- | --- | --- | --- | --- | --- |
| O2B provider | CPHD | HealthPort | Sanrai | FREO2 | AFHIA |
| Start of pilot | April 2023 | October 2023 | June 2024 | February 2024 | June 2023 |
| Facility levels* | Primary<br>Secondary | Primary<br>Secondary | Secondary | Primary<br>Secondary | Primary<br>Secondary |
| Target level for the service | Whole facility | Whole facility | Whole facility | Neonatal and paediatric wards | Whole facility |
| Number of facilities in the pilot | 12 | 7 | 19 | 37 | 5 |
| Number of facilities in the evaluation | 5 | 7 | 5 | 6 | 5 |

\*Facility levels were classified as primary (level 1), secondary (level 2) or tertiary (level 3), based on the Lancet Global Health Commission on Medical Oxygen Security and WHO Emergency and Essential Surgical Care definitions.<sup>1 10</sup>

### Participants and sampling

A sub-set of the facilities which took part in the O2B pilots were purposefully selected for the study (**Table 1**). For the quantitative audits, we planned a minimum of 5 facilities in each country, with input from the O2B providers to select facilities with the most variety in terms of numbers of beds, geographical location, levels of care, duration of O2B pilot implementation, and pre-existing oxygen systems.

For the audits, we aimed to include all ward areas. However, for larger secondary or tertiary hospitals it was not always feasible to complete all ward areas in the limited time for data collection. In these cases, data collectors prioritised high-acuity wards areas (e.g. intensive care, surgical, and general wards), where oxygen is expected to be present for patient care, and de-prioritised low-acuity areas (e.g. outpatients). All facility managers were interviewed for the general facility audit, or a designated manager if they were unavailable. For HCW surveys, all HCWs were eligible to take part, and we used convenience sampling to recruit from those on duty at the time of audit.

For the qualitative interviews, we selected two of the facilities per country where study audits had been done. The facilities with the most variation were purposefully chosen, to try and maximise different experiences across facility contexts. The facility manager interviewed as a key informant, and 2-3 HCWs per facility were selected using convenience sampling. In facilities where O2B was being delivered only to specific wards, staff from these wards were selected. We aimed to have different cadres from each facility represented.

### Quantitative data collection

Data collection teams of 2-3 members were engaged by the O2B providers for the purposes of data collection and were intended to include one biomedical engineer and one person with a clinical background. Data collection at each facility involved: 1) an interviewer-administered general facility questionnaire with the facility in-charge, 2) oxygen equipment audit conducted by the study enumerator with biomedical engineering capacity, 3) an interviewer-administered HCW knowledge survey and structured observation of pulse oximetry measurements.^11^ Training was done by two members of the research team in each country, consisting of 1-day classroom-based, 1-day pilot and a half-day debrief and recap. Data collection teams had regular contact with the research team to clarify any issues. Data checks were run by the research team, and follow-up field checks were conducted in country. Data was collected and managed using REDcap on Karolinska Institutet’s server.^12,13^

### Qualitative interviews

Qualitative interviews were done by independent qualitative researchers local to each setting (AAB, JN, PM, DB, EM), following a semi-structured topic guide (**Supplemental appendix 2**). A generic topic guide was developed and then adapted to each country by the local qualitative researcher following a standardised online training. Interviews were conducted in a convenient private location at the facilities, and were audio recorded, and transcribed verbatim and translated to English (where necessary), by the researcher who conducted the interview. We did not conduct any repeat interviews or return transcripts to participants, and the researchers were not known to the participants.

### Analysis

Quantitative data were summarised using means, medians, and percentages. We used chi^2^ tests to test for differences in oxygen equipment functionality (concentrators, pulse oximeters and cylinders) between those provided as part of the O2B service package and those otherwise available on the wards. We used t-tests to test for differences between HCW knowledge and skills, according to whether they had received O2B provider trainings. Data management and analysis was conducted using Stata v14.

Qualitative data were analysed using Gale et al’s pragmatic framework approach.^14^ During the initial set up of the codebook, 7 transcripts were double coded by CB and PM to ensure consistency in understanding and synthesis of the codes. The initial codebook was reviewed by CK and FK. After agreement was reached, all interviews were coded by PM and CB, and any disagreements discussed with CK. The preliminary analysis was shared with the local qualitative researchers, and comments and feedback incorporated before finalising themes. Analysis was done using NVIVO Version 15.0.

Qualitative and quantitative data were analysed separately and triangulated under common deductive themes in the results. The overall design of the study, and the themes presented in the results were informed by two implementation science frameworks – Proctor’s framework of implementation outcomes and the Consolidated Framework for Implementation Research.^15 16^

### Ethics

The facility in-charge provided permission for audits. All participants for HCW surveys and qualitative interviews provided informed written consent. Ethical approvals were obtained from ethical review boards in each country: Maseno University Scientific and Ethics Review Committee, Kenya (reference: MSU/DRPI/MUSERC/01361/23); National Commission for Science, Technology and Innovation, Kenya (license number: NACOSTI/P/24/37427, reference: #673262); Health Research and Ethics Committee, LASUTH, Lagos State, Nigeria (reference: LREC/06/10/2498); Health Research Ethics Committee, Ogun State, Nigeria (reference: OGHREC/467/2024/290/APP); National Institute for Medical Research, Tanzania (reference: NIMR/HQ/R.8a/Vol.IX/4743); Ugandan National Council for Science and Technology, Uganda (reference: HS5437ES); Makerere University School of Health Sciences REC, Uganda(reference: MAKSHSREC-2023-600) and the Swedish Ethics Review Authority granted approval to process personal data (reference: DNR 2024-00868-01).

### Patient and public involvement

Patients and the public were not involved in the design or execution of the study. The funder was involved in the conceptualisation of the research questions, but took no part in data collection, analysis and the decision to publish.

## Results

### Facilities and participants

Overall, 28/80 facilities were included in the study audits, with data collected from 141 wards. In India, all facilities were secondary-level government hospitals without ICU or surgical capacity, where patients receive oxygen services for free (**Table 2**). Other countries had a mix of facility levels and ownership. In Nigeria, all facilities charged patients for oxygen services, regardless of ownership, while Kenya, Tanzania and Uganda had a mix of free and out-of-pocket provision. All facilities had either outpatient and/or stabilisation rooms, but the extent of inpatient wards varied, with 20/28 having paediatric wards, 12/28 a NICU, 5/28 an ICU, and 17/28 providing surgical services. We recruited 179 HCWs for the survey, with the majority being nurses and midwives (81.6%), 72.1% were female and the median duration of work experience was 6 years (IQR: 2 – 10) (**Supplemental table 2**). For the qualitative interviews, 59 participants from 20 facilities took part (**Supplemental table 3**).

**Table 2:** Characteristics of the health facilities included in the audit.

| Characteristic |  | Kenya<br>(n=5) | Nigeria<br>(n=7) | India<br>(n=5) | Tanzania<br>(n=6) | Uganda<br>(n=5) |
| --- | --- | --- | --- | --- | --- | --- |
| Level of care* | Primary | 3/5 | 3/7 | - | 3/6 | 2/5 |
|  | Secondary | 2/5 | 4/7 | 5/5 | 3/6 | 3/5 |
| Ownership | Public | 1/5 | 2/7 | 5/5 | 4/6 | 1/5 |
|  | Private for profit | 2/5 | 5/7 | - | - | 1/5 |
|  | Private not for profit** | 2/5 | - | - | 2/6 | 3/5 |
| Wards | Outpatient | 5/5 | 6/7 | 3/5 | 6/6 | 5/5 |
|  | Stabilisation room | 5/5 | 6/7 | 5/5 | 3/6 | 3/5 |
|  | Paediatric ward | 4/5 | 5/7 | 2/5 | 6/6 | 3/5 |
|  | Neonatal ward | 2/5 | 4/7 | - | 5/6 | 1/5 |
|  | Adult ward | 4/5 | 6/7 | 1/5 | 6/6 | 3/5 |
|  | HDU/ICU | - | 1/7 | - | 3/6 | 1/5 |
|  | Maternity ward | 4/5 | 5/7 | 5/5 | 6/6 | 3/5 |
|  | General ward | 2/5 | 6/7 | 3/5 | 6/6 | 3/5 |
|  | Surgical ward | 2/5 | 6/7 | - | 6/6 | 3/5 |
| Patients pay for oxygen services | Yes | 3/5 | 7/7 | - | 5/6 | 3/5 |
|  | No | 2/5 | - | 5/5 | 1/6 | 2/5 |
| Financial autonomy of oxygen procurement | Low | 1/5 | - | 1/5 | - | - |
|  | Medium | - | 1/7 | 3/5 | 4/6 | 1/5 |
|  | High | 4/5 | 6/7 | 1/5 | 2/6 | 4/5 |
| Type of oxygen sources available before O2B pilots*** | No | 2/5 | - | - | - | 1/5 |
|  | Oxygen concentrator(s) | 2/5 | 6/7 | 4/5 | 5/6 | 4/5 |
|  | Oxygen cylinders sourced externally | 3/5 | 7/7 | 5/5 | 6/6 | 2/5 |
|  | Piped oxygen from on-site PSA | - | - | 1/5 | - | - |
| Number of oxygen sources available before O2B pilots | None | 2/5 | - | - | - | 1/5 |
|  | Single oxygen source | 1/5 | 1/7 | 1/5 | 1/6 | 2/5 |
|  | Multiple oxygen sources | 2/5 | 6/7 | 4/5 | 5/6 | 2/5 |

\*Facility levels were classified as primary (level 1), secondary (level 2) or tertiary (level 3), based on the Lancet Global Health Commission on Medical Oxygen Security and WHO Emergency and Essential Surgical Care definitions.<sup>1 10</sup>
\*Including faith based facilities
\*\*\*No facilities reported on-site filling of cylinders from a PSA or liquid oxygen

**Table 2:**
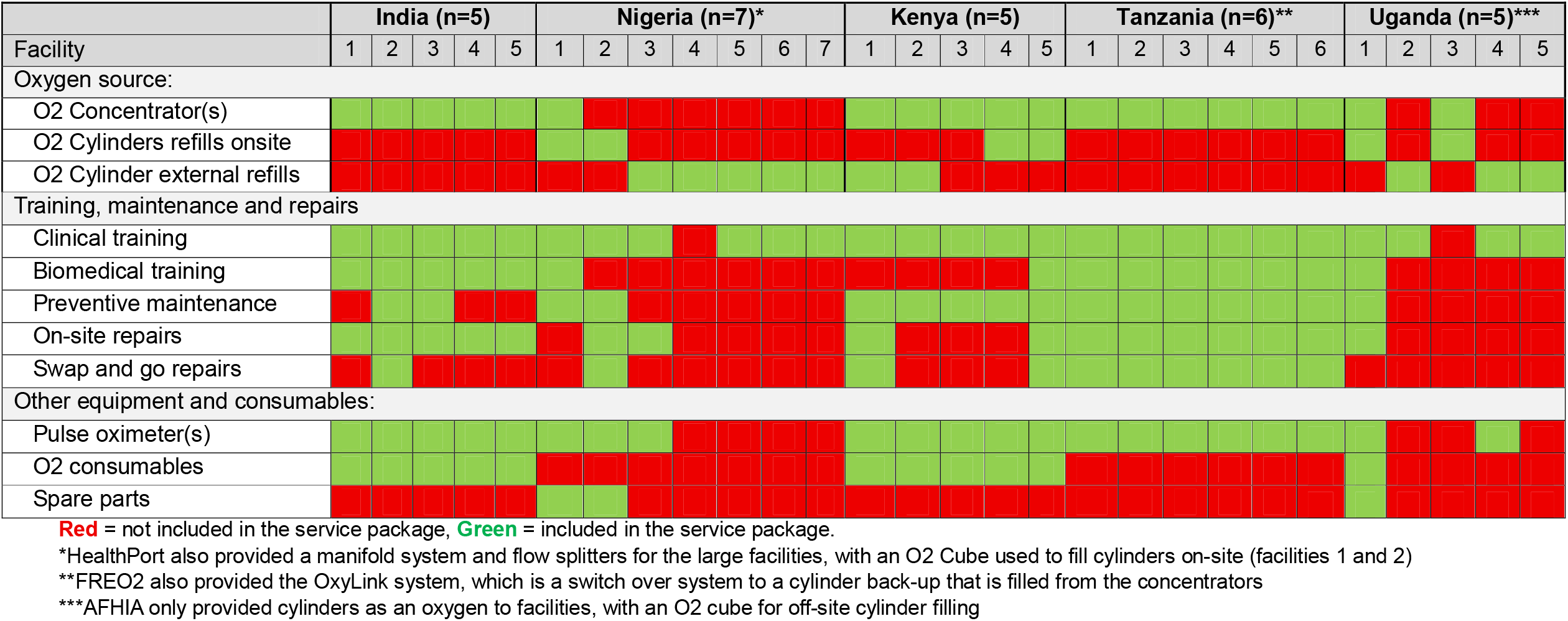
Facility in-charge reported components of the O2B service package from the facility audit questionnaire.

### Defining the O2B package

The O2B packages varied among the O2B providers, and there were variations in what was reported by facilities in-charges serviced by the same O2B provider. Overall, there were apparent gaps in understanding of the O2B package and misalignment between what was provided and reported (see **Supplemental appendix 1** for a description of the O2B packages according to O2B providers).

**Table 2** summarises the different O2B packages, according to the facility in-charge. In all facilities, the O2B package was reported as intended to benefit the “whole facility” (all ward areas and patient groups), except Tanzania where FREO2’s system was designed specifically for neonatal and paediatric care. During the audit, almost all assessed wards in Nigeria had O2B supplied oxygen equipment (55/56, 98%), but this was lower in India (6/13, 46%), Kenya (8/22, 36%) and Uganda (15/21, 71%) (**Supplementary Table 2**). The most common oxygen source included in the O2B package was concentrators (19/28 facilities), and 6/6 facilities from Tanzania and 2/5 from Kenya reported the provision of both concentrators and back-up cylinders. The most common ‘add-ons’ in the packages were: training (26/28 facilities), pulse oximeters (21/28) and preventive maintenance (16/28). In Uganda and Nigeria, solar powered ‘O2 Cubes’ (a 10LPM concentrator with cylinder-filling station) were intended as the main oxygen source. In both cases, the O2B providers sourced cylinders from external suppliers to supplement their service offering.

From the qualitative interviews, there was inconsistency amongst interviewees in terms of what they thought the O2B providers were offering in their package. All interviewees confirmed that provision of an oxygen source was part of the package. However, there were conflicting responses regarding the add-ons, and particularly whether pulse oximeters were provided. Most interviewees stated that maintenance and repair services were included, but experiences of this differed between participants. Some reported prompt and efficient maintenance, with O2B staff available 24/7, while others said they experienced delays – despite a ‘swap and go’ (i.e. the broken equipment is replaced until the repair is completed) service being in place.

> *“It’s been a month since it was sent for repairs, when they took it, we expressed our concerns and asked them to return it as soon as possible”. India, pharmacist*

> *“Whenever there is a need we call him. For example, if we need a mask for an infant, he brings it to us” India, pharmacist*

Interviewee reports on training provided by O2B providers were also inconsistent. Across all five countries, some interviewees reported participating in the training offered by the O2B providers, while others did not, and many recommended repeat or refresher training sessions be conducted to account for high rates of staff turnover

> *“We haven’t received any training so far*.*” Tanzania, facility in-charge*

> *“Training they came like how many times! The first time they came like 2 times*.*” Tanzania, nurse*

### Gaps in oxygen service provision before the O2B pilots

We found mixed gaps in oxygen needs preceding the O2B pilots from both the quantitative and qualitative data. According to the facility in-charge, only three facilities had no oxygen source before the O2B pilots (Kenya=2, Uganda=1, **Table 2**), and 19/28 (68%) already had multiple sources of oxygen, with both externally sourced cylinders (24/28) and oxygen concentrators (21/28) common. However, the qualitative interviews highlighted several challenges with pre-existing oxygen equipment and oxygen supplies.

Interviewees from facilities of different sizes in Tanzania, Nigeria and Uganda reported their pre-existing oxygen equipment was insufficient to meet oxygen demands, with competition for equipment both within and between wards.

> *“You might find that a child needs oxygen and you can’t go and remove oxygen from another patient and put it on this one. There we were losing so many children” Tanzania, nurse*

However, interviewees from secondary facilities in Nigeria and India, reported have adequate oxygen equipment prior to the O2B pilots. In India particularly, interviewees recounted good availability of oxygen concentrators, a result of deliveries by the regional government in response to the COVID-19 pandemic, and oxygen cylinders, which were refilled through a government-run scheme.

> *“We have all kinds of oxygen like small cylinders and jumbo cylinders, there are lot of oxygen concentrators which have been provided by some NGOs and medical facilities also” India, facility in-charge*

Where cylinders were used prior to the O2B pilots, interviewees commonly reported difficulties with refilling due to slow, unreliable, and fragmented supply chains, resulting in facilities struggling to meet oxygen demand. In several instances, interviewees reported receiving cylinders that were half-filled, attributed to leakages during transportation or concerns that suppliers were knowingly supplying half-filled cylinders. Transportation of cylinders was a significant challenge, with many facilities lacking vehicles to transport cylinders, and long distances resulting in high costs:

> *“It is not reliable because several times they have filled our cylinders; we arrive with cylinders that are half of it […] a small cylinder we started traveling with a patient, we haven’t even reached [town], it’s already corrupted*.*” Tanzania, facility in-charge*

> *“The [Chief Medical Officer] office would handle the refiling and return the filled cylinders to the respective [community health centres]. This process was time consuming and required significant attention and care*.*” India, CMO office staff*

Where concentrators were used prior to the O2B pilots, interviewees highlighted unstable electricity/power supplies and unreliable maintenance, leading to equipment down-time and interruptions to oxygen availability. In most facilities, there were no biomedical technicians on site, and challenges with timely access to technicians stationed at regional centers.

> *“[district biomedical engineers] are irregular and sometimes they come once in a year and twice in a year if they try a lot” Uganda, facility in-charge*

Lack of pulse oximeters, along with consumables (e.g. masks, tubing, prongs) was noted as a regular occurrence prior to the O2B pilots, which had limited the use of oxygen in these facilities. Therefore, while the scale and nature of gaps in oxygen services varied before the pilots, consistently there were areas highlighted where O2B providers could improve services.

### Impact of O2B pilots on oxygen availability at the bedside

Generally, the O2B pilots were reported to have been successful in improving medical oxygen availability at the beneficiary health facilities. The quantitative assessment confirmed the improved functionality and appropriateness of the oxygen equipment provided by O2B providers, explaining improved oxygen supply on wards, and therefore patient access.

Overall, 40.0% (159/394) of cylinders on wards were empty, and this was more common for non-O2B cylinders (49.0% versus 30.1%, p-value<0.001). O2B cylinders were more often deemed usable (64.5% versus 40.9%, p<0.001) – **Table 3**. For concentrators 95.0% (38/40) of the O2B provided concentrators were functional, compared to 25.7% (n=18/70, p<0.001) of non-O2B concentrators. The most common reason for non-functionality was being unable to turn on – **Figure 1**. For pulse oximeters 84.0% (21/25) of O2B provided devices were functional, relative to 70.0% (49/70, p=0.172) of non-O2B devices. When looking at neonatal and paediatric wards (n=23), only those who had received O2B equipment had age-appropriate oximeters available (44.4% (8/18) vs. 0% (0/5)), and the only flow splitters we observed were supplied by the O2B provider.

**Table 3:** Equipment functionality for O2B and non-O2B supplied equipment.

|  |  | O2B equipment | Non-O2B equipment | p-value |
| --- | --- | --- | --- | --- |
| Number of cylinders assessed |  | 186 | 208 |  |
| Cylinder usable for patient care* | Yes | 120 (64.5%) | 85 (40.9%) | <0.001 |
|  | No | 66 (35.5%) | 123 (59.1%) |  |
| Number of concentrators assessed |  | 41 | 69 |  |
| Concentrator functional** | Yes | 38 (95.0%) | 18 (24.7%) | <0.001 |
|  | No | 2 (5.0%) | 52 (75.3%) |  |
| Number of pulse oximeters assessed |  | 25 | 70 |  |
| Pulse oximeter functional*** | Yes | 21 (84.0%) | 49 (70.0%) | 0.172 |
|  | No | 4 (16.0%) | 21 (30.0%) |  |

\*Defined as a cylinder with oxygen, a regulator and a working valve.
\*\*Defined as a concentrator that could turn on, with a non-fluctuating flow rate at 2LPM and an oxygen concentration >83%, as measured by the data collector on the day of the audit. Note, non-O2B concentrators in Kenya were not tested.
\*\*\*Defined as device that turns on, provides a SpO<sub>2</sub> measurement >95% in a healthy data collector, and no faults noted.

**Figure 1:**
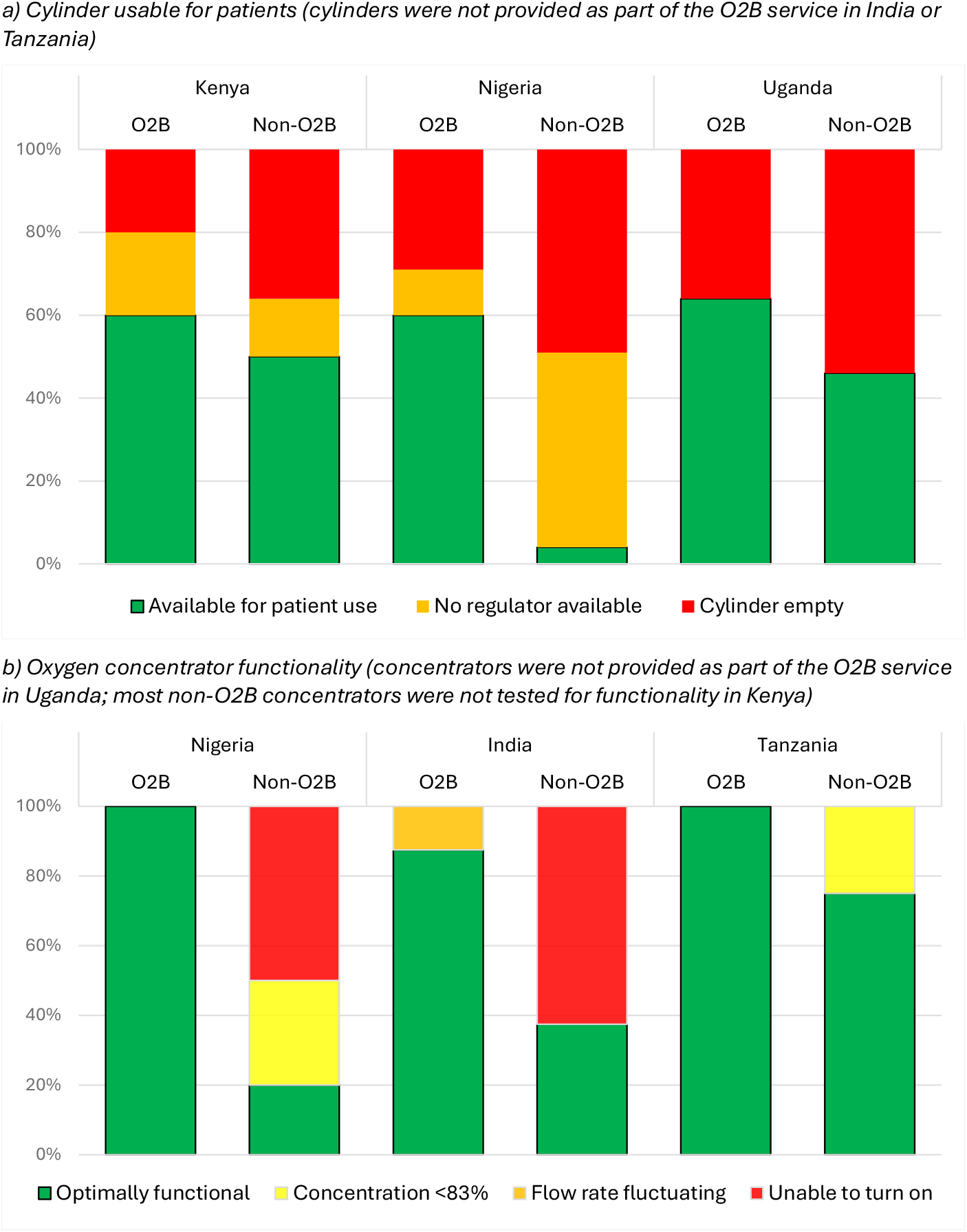
Functionality of equipment by country and source.

Many interviewees reported that O2B pilots had a positive effect on the availability of medical oxygen at the facilities. In particular, the efficiency and quality of oxygen supply from the O2B providers was raised, for example, from Nigeria the guarantee of full oxygen cylinders was described.

> *“For HealthPort, their bottles were bigger and they are fuller” Nigeria, doctor*

The maintenance services provided were reported as more regular and proactive, and in most cases, equipment was replaced when required. Some reported that facility staff also performed routine equipment monitoring and checks and then notified O2B providers when support was required, which led to reductions in equipment downtime.

> *“[it] never took too long to wait for them to come and fix it” Kenya, nurse*

The usability of O2B provided oxygen equipment was also raised as a way in which the pilots improved patient access to oxygen. In particular, the FRE02 Oxylink system in Tanzania was praised for having desirable attributes such as the back-up medical oxygen source in a single package.

> *“It’s easy because it’s always there, you don’t go looking for it. Your job is to take the child there and place them on oxygen” Tanzania, nurse*

> *“They supply one [cylinder] with tap that you just, you know, have to click and then the oxygen is on” Nigeria, doctor*

As a result of O2B providers’ oxygen use monitoring, the interviewees noted that this made it possible to anticipate or forecast future oxygen demand and act to prevent stock-outs. However, requests for additional oxygen sources and pulse oximeters were still made by several interviewees, for example, where O2B equipment was only provided for some wards or pulse oximeters were not part of the service package.

> *“Maybe we get an emergency here, I have to run all the way to maternity and wheel the concentrator to this point” Kenya, nurse*

> *“They need to provide pulse oximeters so that we are able to diagnose hypoxemia” Uganda, nurse*

The improved reliability of oxygen attributed to the O2B pilots was also described as changing patient admissions, with interviewees reporting more patients able to be treated on-site, reducing the need for referrals. Interviewees reported that having oxygen capacity in the facilities improved patient trust in the quality of services. Several linked this to increasing demand for oxygen at the facility, with more critically unwell patients, more deliveries, and more paediatric pneumonia cases.

### Healthcare worker competency, confidence and motivation

In terms of HCW knowledge and practice, several O2B pilots offered staff training to different extents. Shifts in HCW motivation, commitment and confidence in providing oxygen services were apparent to in some contexts. However, these could have been due to reliable availability of oxygen or more convenient and ease to use equipment, than increases in HCW knowledge.

From the HCW surveys, 23.5% (n=42/179) reported being trained in oxygen therapy, and 24.0% (n=43/179) being trained in pulse oximetry, with most of this training reported from O2B providers (22.4%, n=40/179). In terms of knowledge about oxygen therapy, HCWs who had received any form of O2B training performed marginally better (mean score 15.3 versus 13.9 out of 24, p=0.002) – **Table 4**. The difference was due to increased knowledge of pulse oximetry, with some variation between countries (**Figure 2, Supplementary Table 4**).

**Table 4:**
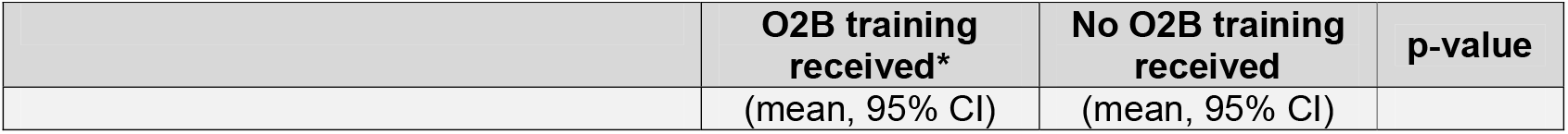
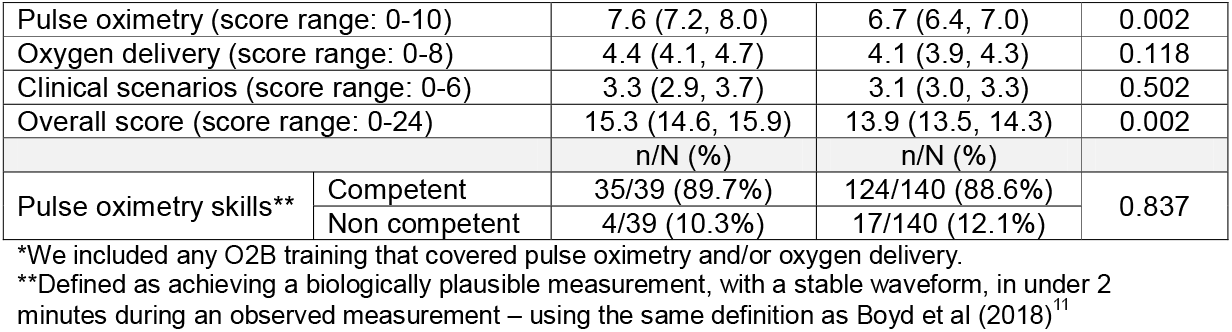
HCW pulse oximetry and oxygen knowledge and skills.

|  | O2B training received* | No O2B training received | p-value |
| --- | --- | --- | --- |
|  | (mean, 95% CI) | (mean, 95% CI) |  |

|  |  |  |  |  |
| --- | --- | --- | --- | --- |
| Pulse oximetry (score range: 0-10) |  | 7.6 (7.2, 8.0) | 6.7 (6.4, 7.0) | 0.002 |
| Oxygen delivery (score range: 0-8) |  | 4.4 (4.1, 4.7) | 4.1 (3.9, 4.3) | 0.118 |
| Clinical scenarios (score range: 0-6) |  | 3.3 (2.9, 3.7) | 3.1 (3.0, 3.3) | 0.502 |
| Overall score (score range: 0-24) |  | 15.3 (14.6, 15.9) | 13.9 (13.5, 14.3) | 0.002 |
|  |  | n/N (%) | n/N (%) |  |
| Pulse oximetry skills** | Competent | 35/39 (89.7%) | 124/140 (88.6%) | 0.837 |
|  | Non competent | 4/39 (10.3%) | 17/140 (12.1%) |  |

\*We included any O2B training that covered pulse oximetry and/or oxygen delivery.
\*\*Defined as achieving a biologically plausible measurement, with a stable waveform, in under 2 minutes during an observed measurement – using the same definition as Boyd et al (2018)<sup>11</sup>

**Figure 2:**
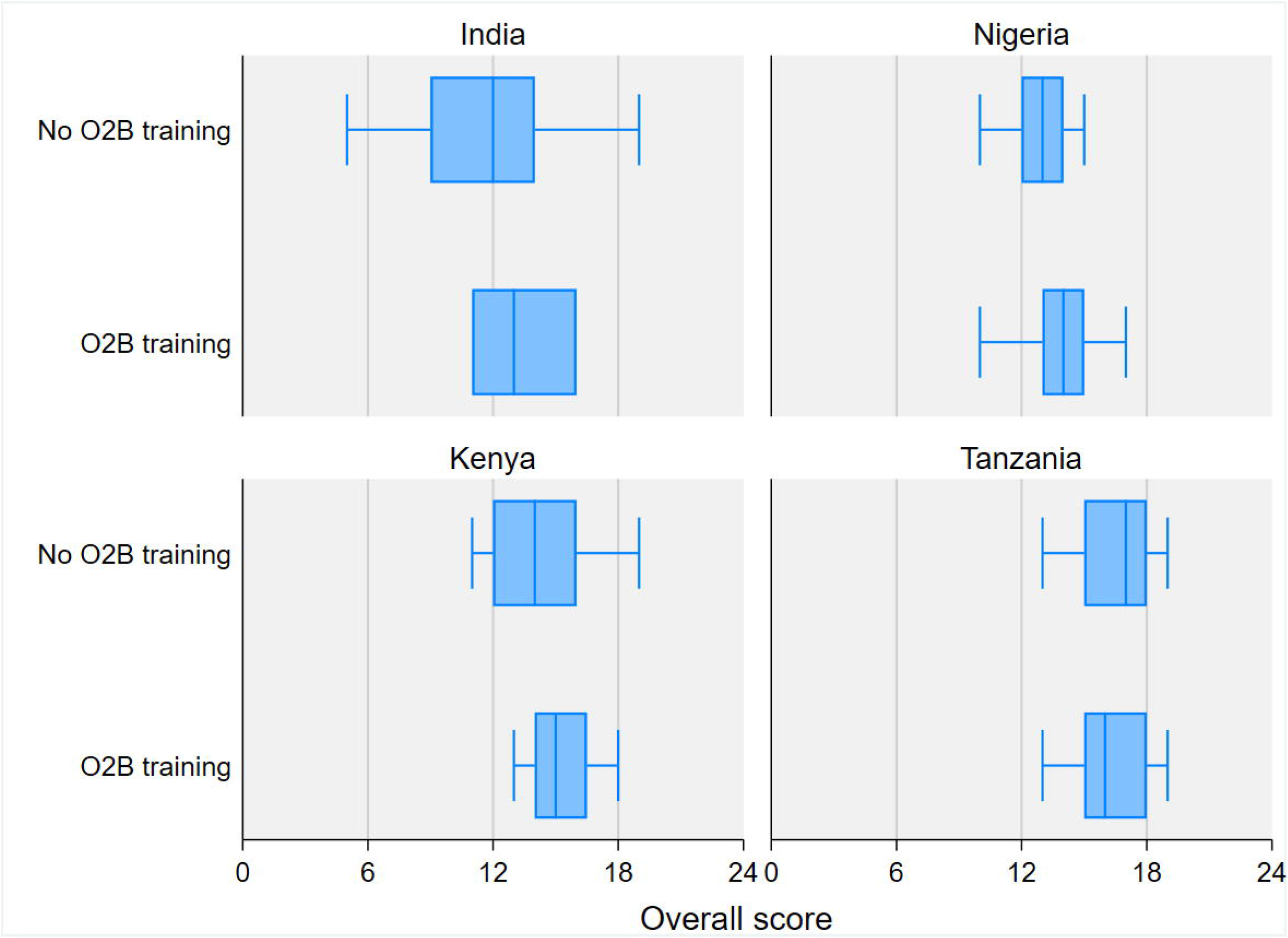
Healthcare worker knowledge by country and O2B provided training. *(training was not part of the O2B service in Uganda)*

From the qualitative interviews in Uganda and Kenya, HCW confidence in oxygen use was lacking before the O2B pilots, given the limited prior exposure to oxygen systems or access to oxygen therapy. Interviewees also commented on their poor knowledge of operating oxygen equipment, which had resulted in broken equipment, inappropriate use and wastage of oxygen. Particularly in Uganda, respondents commented that HCWs had lacked confidence to take responsibility for treating hypoxaemic or critically unwell patients at their facility, preferring to refer them to larger hospitals.

> *“Most people here prefer referring patients without even trying to initiate on oxygen. This is with those people who refused to learn how to give oxygen” Uganda, nurse*

Contrastingly, interviewees in India stated that HCW knowledge and skills were deemed satisfactory before the O2B pilots, given their experience during COVID-19 pandemic. However, this assertion was not supported by the findings from quantitative assessment (**Figure 2, Supplementary table 4**). In Nigeria and Tanzania, interviewees reported that medical oxygen knowledge and skills were considered adequate, especially among staff working on neonatal and paediatric wards.

> *“Almost everybody here knows when to put and when to wean off, even when the doctors are not around” Nigeria, doctor*

Increased staff confidence in providing medical oxygen following the O2B pilot implementation was often reported. Several interviewees commented on the peace of mind that came with improved oxygen availability, and its positive effect as staff took initiative to provide medical oxygen services, even in circumstances where this was previously not an option.

> *“Where this oxygen has helped us a lot, especially us staff, to be confident in performing our duties, because we know that this child, even if she is born with challenges, our oxygen readiness, we are ready with oxygen. So, we know we provide treatment with great confidence unlike before” Tanzania, facility in-charge*

## Discussion

In this implementation evaluation of five unique O2B pilots, we examined whether outsourcing medical oxygen service provision using different approaches improved access to oxygen at the patient bedside. While we were unable to conduct a definitive assessment of patient outcomes, we illustrate important mechanisms through which O2B models appeared to positively change healthcare worker practice and improve patient access to oxygen treatment. However, we also observed a lack of clarity amongst facility staff about what the O2B package contained, and gaps in healthcare worker knowledge and skills remained.

We found that overall, oxygen equipment supplied by the O2B providers was more often functional, available for patient use, and appropriate for the setting (e.g. having age-appropriate pulse oximeter probes). This suggests that the pilots were successful in increasing oxygen access at the patient bedside, and this sentiment was echoed by the qualitative interview participants. One key mechanism in achieving this seemed to be a better design of the medical oxygen system within ward areas (e.g. having a manifold that can support treatment of multiple patients simultaneously). A good example of this was FREO2’s approach in Tanzania for paediatric and neonatal patient that included an oxygen back-up system,^17^ meaning that even when the power went out, the ward could still access oxygen without needing to move cylinders.

While the specific focus of the FREO2 approach on a specific patient group resulted in a tailored and fit-for-purpose approach, there were requests for “whole of facility” facing medical oxygen interventions. There was variation in whether O2B provider’s service package were intended to support the whole facilities oxygen needs or to supplement their existing oxygen supply, with HealthPort in Nigeria representing the other end of this spectrum. HealthPort adapted their service to supply oxygen from multiple sources to meet facility needs, when demand exceeded the production capacity of their O2 Cube, and this ability to adapt was positively reflected with qualitative interviewees. Since this pilot, FREO2 has also expanded their service to a whole facility solution, which is being piloted in the Solomon Islands.^18^ Therefore, a key lesson is not just needing to understand the pre-existing oxygen gaps and systems weaknesses to design a locally appropriate O2B service package, but also ensuring providers have the capacity to adapt.

A second key mechanism that seemed to improve oxygen access was maintenance and repair of concentrators, and quality control of cylinders supplied. Several studies have highlighted the issue of non-functional oxygen equipment on wards,^19^ but there are also examples of how effective preventive maintenance can result in long-term use.^4^ The majority of concentrators classified as non-functional was due to not switching on, rather than supplying low quality oxygen, and therefore were not in active clinical use when audits were conducted. It is possible that the presence of new O2B equipment in facilities replaced rather than supplemented existing equipment, which could lead to deterioration of pre-existing oxygen equipment. For example, in India, interviewees stated that sufficient oxygen supply was not a major challenge before the O2B pilots, but a large proportion of the non-O2B concentrators were not functional. However, we did not collect information on when devices stopped functioning, if repairs had been attempted, or confirmed if devices had been formally decommissioned but left on wards. Given known issue of medical equipment graveyards and limited biomedical engineering workforce capacity,^20 21^ there could be an opportunity for O2B providers to repair other medical equipment within facilities, and offer decommissioning services that free-up clinical space and allows them to re-purpose components.

An interesting finding was the lack of consistency between what end-users said was included in the O2B service, and what the O2B providers said was included (or not), and to some extent this was seen in all settings. For example, in India, there was a ‘swap and go’ service which meant the facility was not missing a device while repairs were done, despite a respondent raising long repair times as a challenge. In Uganda, some respondents spoke about a previous solar oxygen project,^22^ thinking that it was the same as the O2B pilot. In Tanzania, all facilities received training as part of the O2B service, and even if staff turn-over might explain lower than expected coverage, one facility in charge stated that no training had been provided. So, while interviewees were aware that the O2B providers provided a service package, there was generally a lack of appreciation for what that package contained. For frontline staff, while this might be expected (i.e. if oxygen is reliably available, understanding who the supplier is may not be as important), it also presents an opportunity to increase awareness and therefore uptake of add-on services, like training and maintenance.

The third mechanism that seemed to improve oxygen provision was increased healthcare confidence, linked to a reliable oxygen supply. However, there was a clear gap still in healthcare worker knowledge and skills, and while HCWs who had received O2B training did have better knowledge, this difference was marginal and still relatively low. High staff turnovers can be a challenge, and so O2B providers could act as ‘oxygen mentors’, providing frequent on the job training in both their equipment specifically, but also oxygen therapy more broadly. Further work is needed to develop and evaluate how this could be effectively implemented, and whether it impacts on not only knowledge, but also quality of care. O2B providers should also have a motivation to improve HCW capacity, as increased HCW confidence in their capacity to deliver oxygen, when combined with a reliable supply, should result in increased patient use – and therefore, a stronger business case. However, the presence of these feedback loops in the system, can also make it hard for the O2B provider to forecast the oxygen needs in the facility. Systems with local storage capacity (e.g. the LPOS system,^17^ or storing spare cylinders from the O2 Cube) can bring some of this flexibility. Therefore, innovations around oxygen needs tracking, consumption and forecasting,^23^ could lead to more efficient service models.

We had three key limitations. First, we only had data available from one time point after the start of implementation of O2B pilots and there was heterogeneity in length of implementation, components of O2B packages and context across the five study countries. This meant we were unable to draw concrete conclusions about impact or changes over time. We tried to mitigate this by collecting and triangulating different types of data and across settings to increase confidence in our conclusions. We also used convenience sampling for the HCW survey, and most respondents were nurses and midwives, which may not represent the cadre of staff responsible for making oxygen treatment decisions. Secondly, we had different data collectors and qualitative researchers in each country, which may have introduced researcher bias into the data. To try and standardise data collection, the training was led by the same researchers (CB and PM) in all sites, using the same Standard Operating Procedures. However, it’s plausible that some intra-country variations reflect differences in data collection. Third, there may have been social desirability bias, with respondents tailoring their responses if they wanted the O2B pilots to continue. The local qualitative researchers were independent from the O2B providers and were not known to the respondents. However, the quantitative data collectors were employed by the O2B providers, and this may have biased respondents. However, we found both positive and negative sentiments expressed, and differences between facilities, suggesting that a systematic bias was not introduced. Finally, this paper focused on describing if and how the O2b packages improved oxygen access, ignoring cognitive, relational, organizational and team level processes that are known to underlie how change unfolds in such interventions. An accompanying paper aimed to address this limitation in more detail.

Overall, the O2B pilots appeared to have been successful in improving reliable supplies of medical oxygen within the facilities they served. However, given the relatively short periods of implementation, the multiplicity of contexts and cross-sectional nature of the evaluation, additional implementation research and economic evaluation is needed. In particular, understanding which components of service provision packages would be most desired by end users, but also affordable and cost-effective in terms of quality-of-care outcomes.

## Authors contributions

The study was conceived by CK, FEK, MS, PM, CB, HRG and TB, with funding acquisition by CK and FEK. The O2B pilots were led by MN, MH, MR, AA and AR. Protocol and tool development was led by MS, CB and PM, with support from CK and FEK. Qualitative data collection was conducted by DB, AAB, JN, JO, EM and PM, with oversight from CB, and support from FEK and CK. Quantitative data collection was overseen by CB and PM. Qualitative analysis was conducted by CB and PM, with support from CK and FEK. Quantitative data cleaning was done by CB and PM, and analysis was done by PM and CK, with input from FEK. The manuscript was drafted by CK, with support from CB and PM. All authors read and contributed to the final manuscript.

## Acknowledgements

We would like to thank all the healthcare workers and facilities for giving us their time and data and allowing us to discuss their experiences. We would also like to acknowledge the data collectors for their contributions.

## Data availability

Given the qualitative nature of the data, transcripts cannot be sufficiently anonymised for Open Data Access. Data can be requested for the purposes of further academic research, by contacting Carina King and Freddy Eric Kitutu. Requests will be reviewed and discussed with the site qualitative leads, and approval from local ethical committees will need to be sought.

