## Supplemental Appendices for "Pilot implementation of ‘Outsourced Oxygen to the Bedside’ models in five countries: a mixed methods impact assessment"

**Supplementary Appendix 1: Health system and O2B project contexts**

*Kenya*

Kenya has a devolved health system with health services managed at the national and county level. At national level, the Ministry of Health has responsibility over national referral hospitals, is responsible for training and capacity building, health policy and health information systems. At county level, the 47 counties are responsible for primary and secondary care. The level of care spans from Level 1 to level 6 as depicted in table1 The private sector (for profit& not for profit) and other faith based institutions are significant part of the health system.^1^

The O2B pilot in Kenya was led by CPHD, and this evaluation focused on the first phase of implementation in 12 urban facilities in Nairobi. The facilities were a mix of public, private, and faith-based facilities that were selected on a first come first basis. CPHD provided a 5L concentrator and 1 backup cylinder (with a monthly refill), pulse oximeters, a start-up package with consumables, voltage regulator, and cylinder trolley. They also provided add-on services of regular maintenance, corrective maintenance and repair, and provided training on clinical use and maintenance. The O2B package was offered free of charge to facilities at the start of the pilot, with the plan to transition to a paid model.

*Nigeria*

Nigeria has a pluralistic healthcare system, comprising of both the public and private sector. The public health sector is structured into 3 tiers i.e primary, secondary and tertiary with responsibility distributed among the federal, state and local governments. Primary health care is under the jurisdiction of local government areas with facilities including health posts, dispensaries, primary health centres and comprehensive health centres offering basic preventative, promotive and curative services including immunisations, maternal and child health. Secondary care is managed by state ministries of health having general hospitals and some specialist hospitals giving more specialised care. Tertiary health services are managed by the federal government with a jurisdiction over university teaching hospitals, federal medical centres and specialized hospitals offering highly specialized medical services, advanced diagnostics and training for health care professionals.^2^

The O2B pilot in Nigeria was delivered by HealthPort to a larger hospital, and six smaller health centres. Oxygen was delivered using a solar-powered O2 Cube in the hospital, using a manifold and piping, and supplemented with externally sourced cylinders when demand out-weighed the supply of the O2 Cube. The other smaller facilities were supplied with oxygen cylinders. Facilities were charged for the O2B package from the start of the pilots, with HealthPort committing to meet the facilities oxygen needs. They also provided maintenance of HealthPort owned equipment, access to on-site biomedical support, clinical training and pulse oximeters.

*India*

India has a complex health system and combines both the private and public health sectors. The public health system has three levels with primary health care the 1st level with facilities including sub centres, primary health centres and community health centres offering basic outpatient care, maternal and secondary care. Secondary health care is offered at district and subdistrict hospitals offering specialist consultations, inpatient care and emergency services. Tertiary health care facilities include medical colleges and apex hospitals offering advanced medical treatment and specialized surgeries.^3^

The O2B service was provided by Sanrai in India, who worked in 19 secondary level facilities. Their service package included 5L and 10L concentrators, and pulse oximeters for each facility, without any prescription of where equipment should be deployed in the facilities. Sanrai provided continued support to facilities with how to use the equipment and conducted maintenance and repairs for equipment. The equipment was provided free of charge to facilities.

*Tanzania*

Tanzania operates a decentralized health care system with 5 levels of care. Primary health care services are offered at level 1(village health posts, dispensaries and health centres). District hospitals are situated at level 2 of offering general medical services including inpatient and outpatient acre, maternity services and minor surgeries. At level 3 are regional referral hospitals acting as referral facilities for district hospitals and offering specialized medical services and advanced diagnostics. At level 4 are zonal referrals serving multiple regions and providing higher level specialized care. National hospitals constitute level 5 and offer comprehensive specialized services, research and training.^4^

The O2B service in Tanzania was provided by FREO2, in 37 facilities, with a focus on neonatal and paediatric oxygen solutions. They provided their own fit-for-purpose concentrator with improved filtration, indication lights for equipment functionality, voltage stabiliser, and an automatic switch to back-up oxygen cylinders during power-cuts (the OxyLink System). The back-up cylinders were provided by another provider. FREO2 also delivered training and maintenance. The service was provided free to facilities.

*Uganda:*

Uganda has a decentralised health system, with 6 levels of care spanning from village health teams, up to district hospitals, regional and National referral hospitals. The medical oxygen system is complex, involving a mixture of public and private oxygen producers, distributors, and users. The principal governmental and non-governmental players in the oxygen ecosystem are:the Ministry of Health, the Joint Medical Stores, National Medical stores, and National Drug Authority ^5^.

The O2B project in Uganda was led AFHIA and involved the sale of oxygen cylinders to rural health facilities. The cylinders were filled using solar-powered concentrators and compressors (O2 Cube) at a single filling station in rural Uganda (Kambuga). To meet facility demand and while awaiting regulatory approval, additional cylinders were procured from existing oxygen suppliers. Cylinders were distributed and sold to nearby health facilities for a fee (UGX 30,000 = USD $8.57). There was minimal emphasis on oxygen training, add-on technologies such as pulse oximetry, or capacity building for maintenance because the entrepreneurship model focused mainly on supplier-client oxygen relationship with health facilities. Of note, some participating facilities had also participated in past oxygen research projects, involving solar oxygen systems.^6^

**Supplemental Table 1: Description of study locations**

| **Characteristic** | **Kenya** | **Nigeria** | **India** | **Tanzania** | **Uganda** |
| --- | --- | --- | --- | --- | --- |
| Region | Sub-Saharan Africa | Sub-Saharan Africa | South-Asia | Sub-Saharan Africa | Sub-Saharan Africa |
| Income group* | LMIC | LMIC | LMIC | LMIC | LIC |
| GDP(2023, USD)^7^ | 108.04 billion | 363.85 billion | 3.57 trillion | 79.06 billion | 48.77 billion |
| Population (2023) | 55.3 million^8^ | 227.9 million^9^ | 1.4 billion ^10^ | 66.6 million^11^ | 48.7 million^12^ |
| Life expectancy at birth (yrs)^13^ | 66.8 | 63.4 | 67.3 | 66.8 | 66 |
| Under 5 mortality rate (/1000 livebirths)^14^ | 41 | 111 | 29 | 41 | 42 |
| Neonatal mortality rate (/1000 livebirths)^15^ | 22 | 34 | 17 | 21 | 18 |
| Maternal mortality ratio(/100,000 livebirths)^16^ | 379 | 993 | 80 | 276 | 170 |
| Health expenditure (% of GDP) | 4.6%^8^ | 4.8%^9^ | 3.8%^10^ | 3.6%^11^ | 4.7% ^12^ |
| Domestic government health expenditure (%)^17^ | 8.7% | 4.3% | 4.5% | 5.1% | 4.9% |
| Out of pocket expenditure (% of health expenditure)^18^ | 24% | 76% | 46% | 29% | 34% |
| External health expenditure (% of health expenditure)^18^ | 18% | 7% | 1% | 39% | 42% |
| Local definitions for levels of health care | Level of care ranges from community health services(level1) to level 2(dispensaries and clinics) to level 3(health centres) to level4( district hospitals to level 5( county referral hospitals to national referral hospitals^1^ | Primary facilities (health posts, dispensaries, primary and comprehensive health centres), secondary facilities (general, some specialist hospitals), tertiary facilities (university teaching hospitals, federal medical centres, specialized hospitals)^2^ | Three tier levels; 1) Primary health care(sub centres, primary health centres, community health centres), 2) Secondary health care(district and subdistrict hospitals) and 3) tertiary health care (medical colleges and apex hospitals)^3^ | Level 1(village health posts, dispensaries and health centres) to level 2(district hospitals), to level3(regional referral hospitals) to level4(zonal referrals) to level 5(national referrals) ^4^ | Ranges from Village health teams to HCIIs, to HCIIIs, to HCIVs to district hospitals to regional referral hospitals to national referral hospitals ^5^ |

7. WorldBank. World Bank national accounts data, and OECD National Accounts data files, 2025.

8. WHO. World Health Organization 2025 data.who.int, Kenya [Country overview]. 2025

9. WHO. World Health Organization 2025 data.who.int, Nigeria [Country overview]. 2025

10. WHO. World Health Organization 2025 data.who.int, India [Country overview]. 2025

11. WHO. World Health Organization 2025 data.who.int, United Republic of Tanzania [Country overview]. 2025

12. WHO. World Health Organization 2025 data.who.int, Uganda [Country overview]. 2025

13. WHO. World Health Organization 2025 data.who.int, Life expectancy at birth (years), 2024.

14. WHO. World Health Organization Data, Under-five mortality rate, 2024.

15. WHO. World Health Organization 2025 data.who.int, Neonatal mortality rate (per 1000 live births) [Indicator], 2024.

16. WHO. World Health Organization 2025 data.who.int, Maternal mortality ratio (per 100 000 live births), 2025.

17. WHO. World Health Organization 2025 data.who.int, Domestic general government health expenditure (GGHE-D) as percentage of general government expenditure (GGE) (%). 2025.

18. WHO. Global Health Expenditure Database, 2025.

**Supplemental Appendix 2: Key informant semi-structured interview guide (general guide)**

| IDI TOPIC GUIDE  HEALTH CARE PROVIDERS | | | |
| --- | --- | --- | --- |
| Health centre code | Study ID | | Date  [____\|____] / [­____\|____] / ­­[____\|____]  day month year |
| [____\|____] | [____\|____] | |  |
| Position: | | | |
| 1 = In-charge  2 = Senior medical officer  3 = Medical officer  4 = Senior clinical officer | 5 = Clinical officer 6 = Nursing officer  7 = Enrolled nurse  8 = Midwife | 9 = Public health nurse  10 = Nursing aide / assistant  11=District health official | |

| DEMOGRAPHIC INFORMATION | | | | | | |
| --- | --- | --- | --- | --- | --- | --- |
| 1. Age | Years | [____\|____] | 5. Highest level of education or qualification achieved | | | |
| 2. Gender | 1 = Male  2 = Female | [____] | 0 = None  1 = Primary (P1 — P7)  2 = Secondary (S1 — S6)  3 = Certificate | | 4 = Diploma  5 = Bachelor’s degree  88 = Don’t know  99 = Refused to answer | |
| 3. Originally from this area? | 1 = Yes  2 = No | [____] |  |  |  |  |
|  |  |  | 77 = Other____________________________ | | | [____\|____] |
| 4. Number of years worked in this job | | [____\|____] |  | | |  |

| PART 1: INTRODUCTION | | |
| --- | --- | --- |
| Conduct the interview according to the directions below and record information as indicated. | | |
| **Introduction to in-depth interview**  “My name is …………………………………………………………….and I am a researcher for a study with a title **“Exploring approaches to oxygen-as-a-service”** being funded by the Oxygen CoLab, part of Better Futures CoLab and conducted by a team comprising of members from Karolinska Instituet, Makerere University and University of Melbourne. The aim of our study is to explore if external **oxygen as a service (O2aaS)** models are a viable and sustainable solution to increase access to high-quality oxygen delivery. O2aaS is a package of services that can include the oxygen delivery, equipment maintenance, and provider training, provided to healthcare facilities through an external entity. Various O2aas models have been implemented in 5 countries (Kenya, India, Tanzania, Uganda and Nigeria) to improve oxygen access at health facilities. We aim to find out whether the model used by the **grantee (XXXX)** had effect in terms of improving oxygen access to the patients seeking care at the facilities. We have invited you since you have been involved (as a health care worker, facility in charge or district health officer ) at some stage when **the grantee (XXXX)** was implementing its intervention in the facilities. We would want to know your experiences and perception on the oxygen service model/ intervention(s) delivered by the grantee (CPHD, AFHIA, IC Change, Health port and Sanrai) in terms of value added, limitations and whether its sustainable. We hope you will have time to spend with us now to complete this interview. The interview will last for about 30 to 45 minutes and, if you agree, a recording will be made of the discussion such that we record what you say accurately. If you wish to say anything ‘off the record’ this is fine, please indicate to me. The audiotape will only be used by the study team: no one else will hear your voice. It will be kept for **2 years** in our records and will then be erased or destroyed. We are not writing down your names here and no one will be able to identify you in any reports arising out of this research. All records of this interview will be kept securely. Your answers will be looked at together with those of many other health care providers from different facilities and you will not be identifiable in any reports that are published.  Do you have any questions? Do you agree to continue before we start? | | |
| PART 2: O2aas experience, effect and sustainability questions. | | |
| Domains, topic questions, and probes: Use the table below to help you administer the questions during the interview. | | |
| Domain | Topic and Probes | |
| 1. Your role in oxygen therapy | a) Tell me a bit about yourself (prompts: what is your role, how long have you worked in that role). | |
|  | b) Please tell me a bit about how you are involved in medical oxygen in your day-to-day work (prompts: procurement, distribution, clinical use). | |
|  | c) Can you describe the main challenges you face in your role around oxygen? (prompts: supply that meets demand, costs, reliability of oxygen provider, broken equipment). | |
| 2. Awareness of O2aaS | a) What do you know about XXXX’s services? (prompts: How long has XXX been providing oxygen services in your facility/district, when and where did you first here about XXXX oxygen services? | |
|  | b) Can you tell me about XXXX oxygen services? What oxygen commodities were they delivering to the facilities? (Prompts; Did the XXXX oxygen package include equipment (concentrator, cylinders, pulse oximeter, oxygen splitter?) maintenance of equipment? Training of health workers regarding oxygen therapy? Consumables?) | |
|  | c) Are there any aspects of oxygen services that the XXXX oxygen service doesn’t provide that would be useful in your facility? | |
| 3. Experience of O2aaS | a) Can you tell me about ways in which your facility/district procured oxygen before XXXX? (Prompts: equipment, training of staff, maintenance) b) What aspects of O2 procurement were easy before XXXX came in? c) What aspects of O2 procurement were challenging before XXXX came in? | |
|  | d) Have you noticed any differences after changing to the XXXX oxygen service (prompts; What has improved? What has not worked well?) | |
| 4. Context of facilities | **(The questions of this section are to be asked to health care workers and facility managers only)**  a) Can you give me a brief overview of your facility? (Prompts; staff, age, physical layout, size). | |
|  | b) Which physical features of your facility impact/affect the operation of oxygen services? (prompts: lack of ramps/stairs, lack of sockets/power by patient beds, leaking pipes. What factors make oxygen services work well? What factors prevent oxygen services working well?) c) Which organizational features of your facility impact/ affect the operation of oxygen services? (prompts: lack of biomedical staff, responsibility for who orders or notifies stockouts, workplace culture) | |
|  | d) Were any changes needed at your facility to accommodate the XXXX oxygen service? Are any further changes needed? (prompts: physical environment, financing, record keeping, other). | |
| 5. Impact of O2aas | **In this section, we want to know your honest reflections on how well XXXX are meeting oxygen service needs in this facility/district**   a) In your perspective, has anything changed for patients as a result XXXX oxygen services? (Prompts: types of patients, treatments, admissions, referrals) | |
|  | b) What are the most significant benefits of the XXXX oxygen services/model? (Prompts: helpful aspects, benefits for facility staff, usability, ease) | |
| 6. Sustainability of the O2aas model | a) What do you think could affect the XXXX oxygen service continuing in the long term at your facility? (Prompts: cost, usability, facility infrastructure, resources).  b) Is there another oxygen delivery model/intervention to deliver oxygen services in hospitals that you think could work better than the one of XXXX in your facility(prompts; Can you describe that model? Why would you prefer it to the XXXX model?)  c) How important do you think innovations to improve medical oxygen are, compared to other priorities in your hospital? Prompts: what changes are most needed in your facility?  d) If you could change aspects of the XXXX medical oxygen service, what would you change, and why? (Prompts: Do you think you would be able to make these changes? Why / why not?)  **The sustainability questions below should be asked to district managers / policy makers only**   a) Can you give me a brief overview of your district? (prompts: how many people, how many facilities, how many hospitals) Do you have any oxygen production plants in the district (e.g. PSAs)? Is there a competitive oxygen market in this setting? b) What kind of policies, regulations or guidelines did you need to have in place to enable the XXXX oxygen service? (prompts: were new approaches needed, would they be needed if it was a paying service, who has the authority) c) What kind of financial considerations influenced the decision to implement XXXX service pilot? i) What evidence would you need to pay for this going forward? ii) Would you be willing to pay for the XXXX oxygen service if it was offered? iii) What type of payment structure would be feasible? (prompts: service contract, PPP, volume guarantee, service-level agreement) iv) Is there anything [other than cost] that could affect XXXX oxygen service continuing in the long term in your district? (prompts: usability, facility infrastructure, resources) d) How important do you think innovations to improve medical oxygen are, compared to other priorities in your district? Prompts: what changes are most needed in your hospitals? e) Is there another intervention (in another district) to deliver oxygen services in hospitals that you think could work better than current services?  - Can you describe that intervention? - Why would you prefer it to the XXXX?  f) If you could change aspects of the XXXX medical oxygen service, what would you change, and why? Prompt: Do you think you would be able to make these changes? Why / why not? | |
| PART 3: CONTACT SUMMARY FORM (1) | | |
| Interviewer to complete this form after the interview | | |
| Study ID  [____\|____] | | Date  [____\|____] / [­____\|____] / ­­[____\|____]  day month year |
| 1. How would you describe the atmosphere and context of the interview (Include interview location and how this may have affected responses)?  2. What were the main points made by the respondent during this interview?  3. What new information did you gain through this interview compared to previous interviews?  4. Was there anything surprising to you personally? Or that made you think differently?  5. What messages did you take from this interview to improve the CPHD oxygen delivery model?  6. Were there any problems with the topic guide (e.g. wording, order of topics, missing topics) you experienced in this interview? | | |

**Supplemental Table 2: Facility and healthcare worker participants**

|  | **Kenya** | **Nigeria** | **India** | **Tanzania** | **Uganda** |
| --- | --- | --- | --- | --- | --- |
| Healthcare worker knowledge and skills | | | | | |
| Number of HCWs | 31 | 28 | 15 | 34 | 71 |
| Doctor | 1 (3.2%) | - | 3 (20.0%) | 4 (11.8%) | 6 (8.5%) |
| Clinical officer | 10 (32.3%) | - | - | 1 (2.9%) | 4 (5.6%) |
| Nurse/midwife | 19 (61.3%) | 28 (100%) | 10 (66.7%) | 29 (85.3%) | 60 (84.5%) |
| Other | 1 (3.2%) | - | 2 (13.3%) | - | 1 (1.4%) |
| Gender |  |  |  |  |  |
| Male | 9 (29.0%) | 3 (10.7%) | 2 (13.3%) | 9 (26.5%) | 27 (38.0% |
| Female | 22 (71.0%) | 25 (89.3%) | 13 (86.7%) | 25 (73.5%) | 44 (62.0%) |
| Years experience (median, IQR) | 4 (2 – 8) | 9 (3.5 – 11) | 7 (5 – 10) | 10 (5 – 15) | 5 (1.5 – 10) |
| Years in facility (median, IQR) | 1 (0.5 – 3) | 3 (1 – 7) | 3 (1 – 10) | 7.5 (2 – 14) | 2 (1 – 6) |
| Ward assessments | | | | | |
| Number of wards assessed | 22 | 56 | 13 | 29 | 21 |
| O2B equipment present | 8 (36.4%) | 55 (98.2%) | 6 (46.2%) | 10 (34.5%) | 15 (71.4%) |
| O2B equipment not present | 14 (63.6%) | 1 (1.8%) | 7 (53.9%) | 19 (65.5%) | 6 (28.6%) |
| Types of wards |  |  |  |  |  |
| OPD / emergency | 6 (27.3%) | 10 (17.9%) | 5 (31.3%) | 6 (20.7%) | 2 (9.5%) |
| Neonatal | 1 (4.6%) | 3 (5.4%) | - | 3 (10.3%) | - |
| Paediatric | 3 (13.6%) | 4 (7.1%) | - | 4 (13.8%) | 5 (23.8%) |
| Adult /general | 5 (22.7%) | 17 (30.4%) | 3 (18.8%) | 10 (34.5%) | 4 (19.1%) |
| Surgery | - | 2 (3.6%) | - | - | 4 (19.1%) |
| Maternity | 5 (22.7%) | 7 (12.5%) | 5 (31.3%) | 6 (20.7%) | 4 (19.1%) |
| Other | 2 (9.1%) | 13 (23.2%) | 3 (18.8%) | - | 2 (9.5%) |
| Equipment assessments | | | | | |
| Cylinders assessed | 33 | 154 | 7 | 37 | 163 |
| O2B provided | 5 (15.2%) | 117 (76.0%) | 0 | 17 (46.0%) | 47 (28.8%) |
| Not O2B provided | 28 (84.9%) | 37 (24.0%) | 7 (100%) | 20 (54.0%) | 116 (71.2%) |
| Concentrators assessed | 22 | 13 | 23 | 29 | 37 |
| O2B provided | 5 (22.7%) | 3 (23.1%) | 15 (65.2%) | 17 (58.6%) | 0 |
| Not O2B provided | 17 (77.3%)* | 10 (76.9%) | 8 (34.8%) | 12 (41.4%) | 37 (100%) |
| Pulse oximeters assessed | 10 | 35 | 6 | 23 | 21 |
| O2B provided | 5 (50.0%) | 8 (22.9%) | 2 (33.3%) | 9 (39.1%) | 0 |
| Not O2B provided | 5 (50.0%) | 27 (77.1%) | 4 (66.7%) | 14 (60.9%) | 21 (100%) |

*14 non-O2B concentrators in Kenya were not assessed during the audit

**Supplemental Table 3: Participants in the qualitative interviews**

|  | **Nigeria** | **India*** | **Tanzania** | **Kenya** | **Uganda** |
| --- | --- | --- | --- | --- | --- |
| **Total** | 16 | 11 | 12 | 9 | 11 |
| **Cadre** |  |  |  |  |  |
| District health officer | - | 1 | 1 | - | - |
| Facility in-charge | 1 | 3 | 2 | 3 | 4 |
| Doctor | 4 | 2 | 2 | - | 1 |
| Nurse / midwife | 3 | - | 5 | 4 | 4 |
| Other clinical staff** | 2 | - | - | 1 | - |
| Pharmacist | - | 5 | - | - | 1 |
| Biomedical / technical staff | 1 | - | 1 | 1 | 1 |
| Administrative staff | 5 | - | 1 | - | - |
| **Gender** |  |  |  |  |  |
| Male | 5 |  | 6 | 4 | 10 |
| Female | 11 |  | 6 | 5 | 1 |

**Supplemental Table 4: Healthcare worker pulse oximetry and oxygen knowledge and skills, according to cadre and work experience (n=179)**

|  | | n | Pulse oximetry knowledge  *Score≥7/10* | O2 concentrator & delivery knowledge  *Score≥6/8* | O2 therapy clinical scenarios  *Score ≥4/6* | Overall knowledge score  *Score≥16/24* | Pulse oximetry skills |
| --- | --- | --- | --- | --- | --- | --- | --- |
| Job title | Doctor* | 14 | 12 (85.7%) | 0 | 8 (57.1%) | 8 (57.1%) | 8 (57.1%) |
|  | Nurse / midwifes | 146 | 93 (63.7%) | 17 (11.6%) | 46 (31.5%) | 46 (31.5%) | 99 (67.8%) |
|  | Clinical officers | 15 | 10 (66.7%) | 1 (6.7%) | 10 (66.7%) | 10 (66.7%) | 9 (60.0%) |
|  | Others | 4 | 1 (25.0%) | 1 (25.0%) | 2 (50%) | 2 (50.0%) | 2 (50%) |
| Number of years worked clinically | <5 | 72 | 54 (75%) | 4 (5.6%) | 25 (34.7%) | 24 (33.3%) | 48 (66.7%) |
|  | 5-10 | 64 | 41 (64.1%) | 9 (14.1%) | 25 (39.1%) | 20 (31.3%) | 42 (65.6%) |
|  | 11-20 | 32 | 25 (78.1%) | 4 (12.5%) | 13 (40.6%) | 13 (40.6%) | 20 (62.5%) |
|  | >20 | 11 | 8 (72.7%) | 2 (18.2%) | 2 (18.2%) | 6 (54.6%) | 8 (72.7%) |
| Country | Kenya | 31 | 23 (74.2%) | 1 (3.2%) | 16 (51.6%) | 16 (51.6%) | 30 (96.8%) |
|  | Nigeria | 28 | 15 (53.6%) | 5 (17.9%) | 5 (17.9%) | 5 (17.9%) | 27 (96.4%) |
|  | India | 15 | 4 (26.7%) | 2 (13.3%) | 4 (26.7%) | 4 (26.7%) | 9 (60.0%) |
|  | Tanzania | 34 | 34 (100%) | 3 (68.8%) | 24 (70.6%) | 24 (70.6%) | 34 (100%) |
|  | Uganda | 71 | 40 (56.3%) | 8 (11.3%) | 17 (23.9%) | 17 (23.9%) | 59 (83.1%) |
| O2B training provided** | Yes | 40 | 35 (87.5%) | 6 (15%) | 19 (47.5%) | 19 (47.5%) | 32 (80.0%) |
|  | No | 139 | 81 (58.3%) | 13 (9.4%) | 47 (33.8%) | 47 (33.8%) | 86 (61.8%) |

*Includes: consultants, SMO, interns

**We considered training as being provided if it covered pulse oximetry and/or oxygen delivery.
